# Modelling the impact of COVID-19-related control programme interruptions on progress towards the WHO 2030 target for soil-transmitted helminths

**DOI:** 10.1101/2020.11.01.20220376

**Authors:** Veronica Malizia, Federica Giardina, Carolin Vegvari, Sumali Bajaj, Kevin McRae-McKee, Roy M Anderson, Sake J de Vlas, Luc E Coffeng

## Abstract

**Background:** On the 1^st^ of April 2020, the World Health Organization (WHO) recommended an interruption of all neglected tropical disease control programmes, including soil-transmitted helminths (STH), in response to the COVID-19 pandemic. This paper investigates the impact of this disruption on the achieved progress towards the WHO 2030 target for STH.

**Methods:** We used two stochastic individual-based models to simulate the impact of missing one or more preventive chemotherapy (PC) rounds in different endemicity settings. We also investigate the extent to which the impact can be lessened by mitigation strategies, such as semi-annual or community-wide PC.

**Results:** Both models show that even without a mitigation strategy, control programmes will catch up by 2030. The catch-up time is limited to a maximum of 4.5 years after the interruption. Mitigations strategies may reduce this catch-up time by up to two years and can even increase the probability of achieving the 2030 target.

**Conclusions:** Though a PC interruption will only temporarily impact the progress towards the WHO 2030 target, programmes are encouraged to restart as soon as possible to minimise the impact on morbidity. The implementation of suitable mitigation strategies can turn the interruption into an opportunity to accelerate the progress toward reaching the target.

## Introduction

Globally, more than one billion people in developing countries are estimated to be infected with at least one species of soil-transmitted-helminths (STH).^1^ The STH species that mainly affect humans are two species of hookworm (*Necator americanus, Ancylostoma duodenale*), roundworm (*Ascaris lumbricoides*), and whipworm (*Trichuris trichiura*). STH represent a major cause of morbidity, particularly in children. Common STH-related morbidities are anaemia, growth impairment, respiratory problems and malnutrition due to malabsorption and nutrient loss. More severe morbidity is usually associated with moderate-to-heavy intensity (M&HI) of parasitic infection.^1^ The control of morbidity drives the definition of the global target for the elimination of STH as a public health problem (EPHP), that the World Health Organization (WHO) has set by 2030. The target is defined as reaching a prevalence of M&HI infections below 2% in school-age children (SAC).^2^ The current guidelines provided by the WHO to achieve this goal recommend preventive chemotherapy (PC) for pre-SAC (age 2-5 years) and SAC (age 5-15 years) once per year in moderate endemicity settings (20-50% pre-control prevalence of any intensity of infection in SAC) and twice per year in high endemicity settings (>50%). No PC is recommended in areas with low pre-control STH prevalence (<20%).^2^

On the 1^st^ of April 2020, the WHO recommended an interruption of all neglected tropical disease control programmes, including STH, in response to the pandemic caused by the novel coronavirus SARS-CoV-2.^3^ In respect of social distancing, the main public health measure taken to contain the spread of SARS-CoV-2, WHO advised that evaluation activities and PC administration should be postponed until further notice. The impact of this disruption on gains achieved in the control of STH thus far, requires investigation. The time control programmes will take to catch-up with the past and predicted progress will have implications on the time (delay) and feasibility of reaching the target by 2030. To minimise the losses, suitable mitigation strategies may have to be implemented when programmes resume.

We use two independently developed stochastic individual-based models to simulate the impact of missing or postponing one or more PC rounds on the control of STH in different endemicity settings, defined by the pre-control situation, and for all three STH species. We compare the scenario without interruption (baseline scenario) with different “interruption scenarios” which include re-starting the treatment after different interruption lengths, with or without plausible mitigation strategies. For each scenario, we express the estimated impact in terms of three measures: i) the catch-up time, i.e. the time after the interruption required for the M&HI prevalence to catch up with the scenario without interruption, ii) the probability to reach the control target set by the WHO, and iii) the delay, i.e. how much longer it takes to reach the target with respect to the baseline scenario. We investigate the extent to which these quantities can be improved by suitable mitigation strategies.

## Materials and Methods

### Transmission models

We used two stochastic individual-based models independently developed by Erasmus MC (EMC) and Imperial College London (ICL).^4,5,6^ These models are used to simulate the process of transmission of STH in an age-structured human population, through an environmental reservoir of infection (eggs/larvae). Within a cycle, human hosts can be infected and contribute to the reservoir.

The life cycle of worms within the human hosts is also modelled. In both models, a single-slide Kato-Katz faecal smear test is simulated, providing egg counts for each individual. The two models have been calibrated in order to reproduce the same endemicity settings at pre-control level, by varying the species-specific parameters regulating the transmission conditions, i.e. the transmission rate (EMC model) or basic reproduction number R_0_ (ICL model), as well as the level of exposure heterogeneity, which indicates the extent of aggregation of worms among hosts. With both models, we simulate the impact of PC, assuming an effective treatment coverage of 75% of the target population (pre-SAC and SAC or the whole community). Individual participation in PC is assumed to be random, meaning that each round a new random fraction of the population participates. The population is treated with albendazole which we assume to kill 94% of hookworm, 99% of *A. lumbricoides* and 60% of *T. trichiura* adult worms.^7^ A complete description of the parameters used in both models to run simulations, is available in **Supplementary Table 1**. All analyses were performed in accordance with the PRIME-NTD criteria^8^ (**Supplementary Table 2**).

### Scenarios and mitigation strategies

Two endemicity settings are considered: namely, moderate transmission (20-50% pre-control prevalence evaluated in SAC by single Kato-Katz) and high transmission (pre-control prevalence > 50%). For each species, we ran 500 repeated simulations with both models for each of the six different control scenarios outlined in **Figure 1**, over a timeline of 12 years (2018-2030) where PC is initiated in 2019. The “No-interruption” scenario (baseline) assumes that no interruption occurs, and the PC continues at the same frequency and coverage until 2030. Then, we consider that in 2020 a disruption due to the COVID-19 pandemic causes one or more rounds of PC to be missed and that the programme is resumed normally, after 6, 12 or 18 months. As represented in **Figure 1**, according to the endemicity setting and the different “interruption scenarios”, the number of rounds lost is one to three. To mitigate the potential progress lost during the interruption of PC, we also consider scenarios where programmes are resumed one year after the interruption (12-month interruption), with the addition of the following mitigation strategies: i) doubling the frequency of PC for the whole period after the interruption, ii) providing community-wide PC (through mass drug administration or MDA) for only one year after the interruption and then going back to treating only pre-SAC and SAC. In the high endemicity settings, we do not model the scenario of doubling the frequency of PC, because we consider the administration of four PC rounds per year unrealistic/infeasible. Also, the additional impact of more than two rounds per year of PC is negligible, given an assumed expected worms lifespan of 1 (*A. lumbricoides)*, 1 (*T. trichiura)*, 2-3 (hookworm) years (see **Supplementary Table 1**).

**Figure 1.**
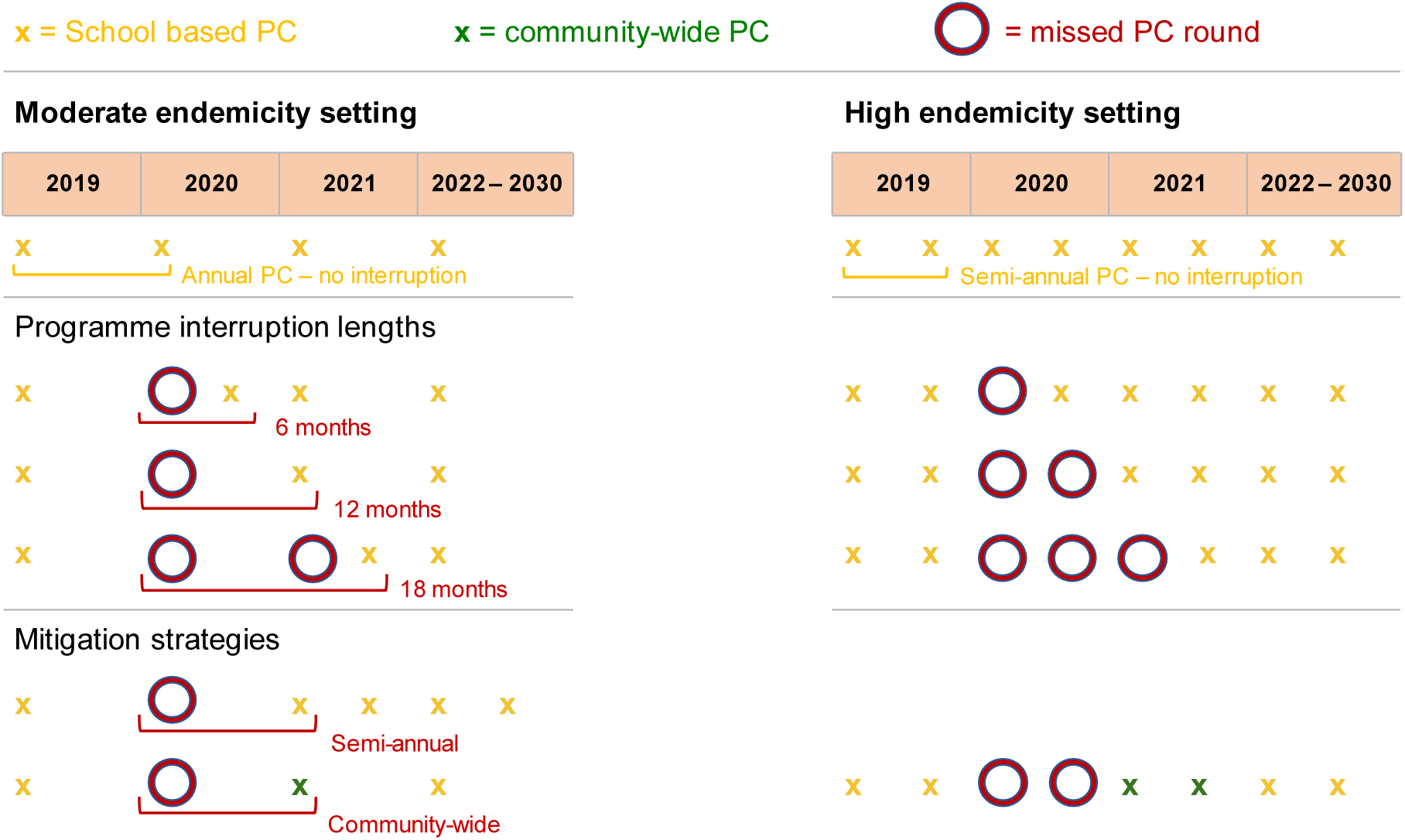
Conceptual diagram explaining the frequency of preventive chemotherapy (PC) for each simulated scenario. School-based PC is distributed once per year in moderate endemicity settings and twice per year in the high endemicity settings. Each cross represents one PC round. Each circle represents one missed round. Each green cross represents one round of community-wide PC. Interruption scenarios without mitigation strategies are presented for different interruption lengths, mitigation strategies are assumed to start 12 months after the interruption.

### Catch-up times and delays

We predict the impact of PC interruption by comparing the dynamics of the prevalence of M&HI infections in SAC in the different simulated scenarios. The results are expressed in terms of three outcome measures: i) the catch-up time, defined as the time from the interruption, until the M&HI prevalence in SAC becomes equal or lower than the one in the scenario without interruption, based on pair-wise differences between single stochastic simulations; ii) the probability to reach the control target set by the WHO (i.e. the proportion of stochastic simulations showing a M&HI prevalence below 2% at time point 2030); and iii) the delay in reaching the target, which is the additional amount of time needed to reach a M&HI prevalence below 2% (that remains below 2% until 2030) with respect to the scenario without interruptions. The delays are computed based on pair-wise differences between single stochastic simulations that reach the target in the baseline scenario.

## Results

### Interruptions without mitigation strategies catch up by 2030

In **Figure 2**, we compare the prevalence of M&HI infection in the baseline scenario with the interruption scenarios without mitigation strategies, in which up to two PC rounds were missed due to a 6, 12 or 18-month programme interruption, starting in 2020. The figure relates to the moderate endemicity setting and it shows that for all species, after interruption the prevalence eventually catch up with the prevalence of the baseline scenario, according to both models. In areas where *A. lumbricoides* is the dominant species it takes slightly longer to recover the progress made by previous control efforts. **Supplementary Figure 1** shows the analogous results for the high endemicity setting, where the M&HI prevalence of the baseline scenario is compared with the interruption scenarios without mitigations, in which up to three PC rounds were missed. The figure shows that for all species, the prevalence in the interrupted scenarios catch up with the prevalence of the baseline scenario by 2030. The highest impact of interruption on the M&HI prevalence is observed for *T. trichiura*, if an interruption of 12 or 18 months occurs. **Table 1** summarises the impact of programme interruptions in terms of estimated catch-up times, i.e. the average time needed for the M&HI prevalence in SAC in the interruption scenarios to catch up with the baseline scenario. The values are expressed in years from the start of the interruption. In moderate prevalence settings, if programmes resume after 6 months, the progress in reaching EPHP will be recovered in less than two years (**Table 1**) and an interruption of 12 months will require around three years in both moderate and high endemicity settings. Interestingly, in moderate endemicity settings an interruption of 18 months does not increase the time needed to catch up with the baseline scenario, with respect to an interruption of 12 months (**Table 1**). **Figure 2** shows that this is due to the beneficial effect of having the first PC round after 18 months of interruption, which happens only six months before the following normal-scheduled round (see **Figure 1** for the scheme of scenarios), effectively resulting in one year of semi-annual treatment. This pattern is not observed in the high endemicity settings, because all the scenarios already include semi-annual PC (**Supplementary Figure 1**). In high endemicity settings, prevalence catch up after different interruption’s lengths within on average 2.5 years (hookworm) and within on average 3.5 years (*A. lumbricoides*), while *T. trichiura* prevalence requires up to 4.5 years according to both models (**Table 1)**.

**Table 1.** Model-predicted time (mean number of years since start of interruption [95%-confidence interval]) required to catch up with the progress towards the 2030 target after 6, 12, or 18 months of programme interruption, compared to a scenario without interruption and assuming that no mitigation strategy is implemented.

| Species | Model | Moderate endemicity setting |  |  | High endemicity setting |  |  |
| --- | --- | --- | --- | --- | --- | --- | --- |
|  |  | 6-month interruption | 12-month interruption | 18-month interruption | 6-month interruption | 12-month interruption | 18-month interruption |
| *A. lumbricoides* | ICL | 1.35 [0.5; 5] | 2.89 [1; 8] | 2.78 [2; 8] | 1.76 [0.5, 4] | 2.69 [1, 6] | 3.65 [2, 8.5] |
|  | EMC | 1.25 [1; 3] | 3.56 [1; 9] | 2.80 [2; 8] | 1.48 [0.5; 4] | 2.14 [1.5; 4] | 2.67 [2; 5] |
| Hookworm | ICL | 1.20 [0.5, 3] | 2.28 [1, 5] | 2.35 [1.5, 5] | 1.12 [0.5, 3] | 1.67 [1, 3.5] | 2.15 [1.5, 4] |
|  | EMC | 1.29 [0.5; 4] | 2.43 [1; 6] | 2.47 [1.5; 6] | 1.34 [0.5; 3.5] | 1.98 [1; 4] | 2.54 [1.5; 5] |
| *T. trichiura* | ICL | 1.32 [0.5, 5] | 2.26 [1, 5] | 2.41 [1.5, 6] | 1.87 [0.5, 4.5] | 3.27 [1.5, 6] | 4.65 [2, 10] |
|  | EMC | 1.50 [0.5; 5] | 2.85 [1; 8] | 2.82 [1.5; 7] | 2.02 [0.5; 5] | 3.31 [1.5; 7.5] | 4.47 [2; 9] |

**Figure 2.**
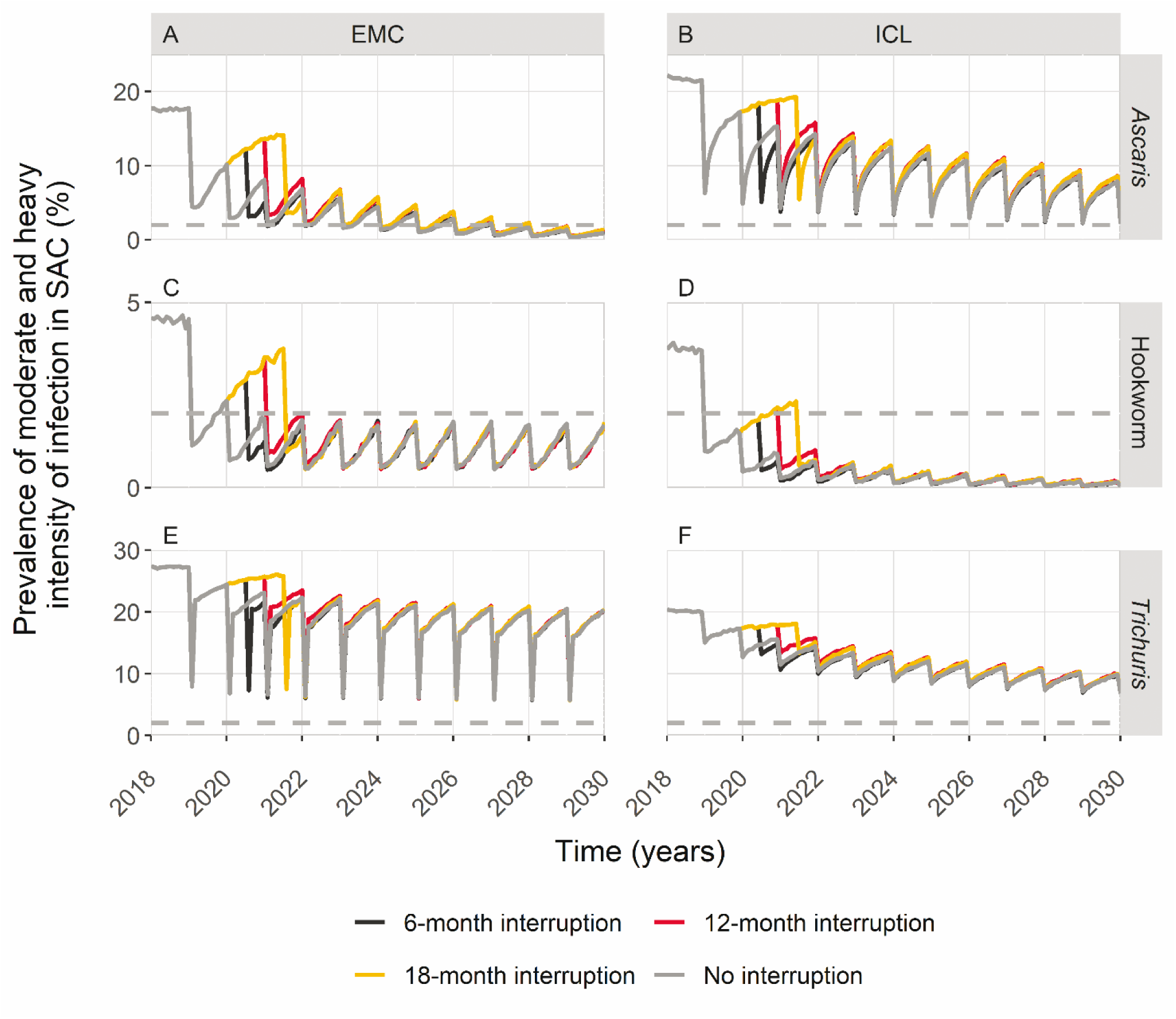
Timeline of moderate-to-heavy intensity infections prevalence in moderate endemicity settings for different interruption lengths if programmes resume without mitigation strategies. The comparison between “no interruption” and restarting after 6, 12 or 18 months is presented by line colours. The horizontal dashed line represents the 2% threshold set by the WHO to assess the goal. Results from both the EMC model (A, C, E panels) and the ICL model (B, D, F panels) are shown, for all STH species.

Interruptions up to 18 months do not have a strong impact on the probability to reach the WHO target for hookworm, and the time to reach the WHO target will be delayed by between 0.34 (95% CI= [-1; 1]) and 0.49 (95% CI= [-2; 2]) (EMC model) and between 0.39 (95% CI = [-1; 1]) and 1.49 (95% CI = [0; 3]) years (ICL model) (**Supplementary Table 3**). A negative value means that the moment when the target is reached is anticipated with respect to the baseline scenario. The M&HI prevalence of *A. lumbricoides* is likely to reach levels below 2% with 83.2% (416/500) probability in the baseline scenario (EMC) and 26.2% (131/500) (ICL) (**Supplementary Table 4**).

Interruptions do not affect the probability of reaching the target according to the EMC model, but they can delay the time when the target is reached by 0.5 [-1; 2] (6-month interruption) to 1.4 [0; 3] years (18-month interruption). According to both models, it is not feasible to reach the target by 2030 even without interruptions in settings where *T. trichiura* is the dominant species, due to the relatively low efficacy of albendazole against this STH infection.

### Mitigation strategies help recover and speed up the progress

The M&HI prevalence dynamics of the scenarios with two mitigation strategies (semi-annual PC and one round of community-wide PC) are compared for the moderate endemicity setting in **Figure 3**. The figure also shows the scenario without any mitigation strategy for comparison. The analogous figure is presented for the high endemicity setting in **Supplementary Figure 2**. We computed the differences between the catch-up time of each mitigation strategy and the catch-up time of the same interruption’s length scenario without mitigations. The values are summarized in **Table 2**. With semi-annual PC when resuming programmes in moderate endemicity settings, the catch-up will be speeded up of less than two years with respect of resuming without mitigation strategies for *A. lumbricoides;* of about one year in the cases of hookworm and *T. trichiura*. If control programmes implement a one-year period of community-wide PC and then revert to targeting SAC and pre-SAC, about one year is gained towards the catch-up for *A. lumbricoides* (both models) and *T. trichiura* (EMC model). In the other cases the time required to catch up with past progress is not significantly shorter than resuming without mitigation strategies. In high endemicity settings, mitigation by means of community-wide PC is beneficial in terms of catch-up time only in the case of *T. trichiura*, allowing the prevalence to catch up about one year sooner than if no mitigation is applied, for both models (**Table 2**).

**Table 2.** Model-predicted differences of time (mean number of years [95%-confidence interval]) required for each mitigation strategy (semi-annual or community-wide preventive chemotherapy (PC)), to catch up with progress towards the 2030 target, compared to the catch-up time required if resuming without mitigation strategies. All values relate to the case of a one-year programme interruption.

| Species | Model | Moderate endemicity setting |  | High endemicity setting |
| --- | --- | --- | --- | --- |
|  |  | Semi-annual PC | Community-wide PC | Community-wide PC |
| *A. lumbricoides* | ICL | 1.26 [0; 6] | 0.77 [-1; 5] | 0.73 [-1; 4] |
|  | EMC | 1.70 [0; 7] | 1.32 [-1; 6] | 0.39 [-0.5; 2.5] |
| Hookworm | ICL | 0.71 [0; 3] | 0.35 [-1; 3] | 0.09 [-1; 1.5] |
|  | EMC | 0.80 [0; 4] | 0.31 [-1.5; 3] | 0.24 [-1; 2] |
| *T. trichiura* | ICL | 0.71 [-1; 4] | 0.17 [-2; 3] | 0.82 [-1; 4] |
|  | EMC | 1.13 [0; 5.5] | 1.05 [0; 6] | 1.79 [0; 6] |

**Figure 3.**
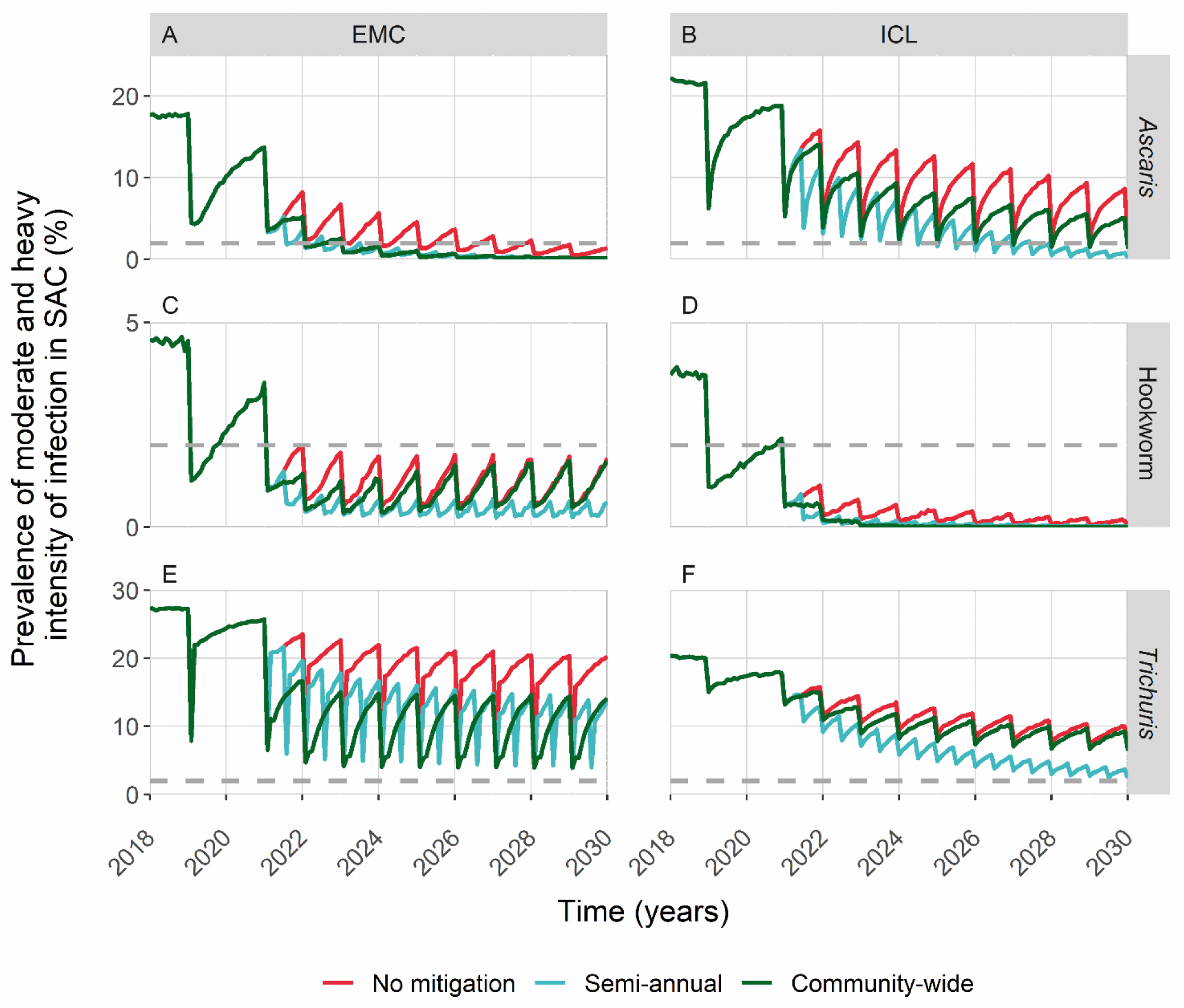
Timeline of moderate-to-heavy intensity infections prevalence in moderate endemicity settings, if programmes resume one year after the interruption, i) without mitigation strategies (red line), ii) doubling the frequency (turquoise line) iii) providing a first year of community-wide MDA before going back to the current WHO treatment guidelines (dark green line). The horizontal dashed line represents the 2% threshold set by the WHO to assess the goal. Results from both the EMC model (A, C, E panels) and the ICL model (B, D, F panels) are shown, for all STH species.

The implementation of mitigation strategies can be essential to enhance the chance to reach the WHO target or to speed up the time to the achievement of the target. The 66.6% (333/500) probability (EMC model) to reach the target, observed for hookworm in the baseline scenario, increases to 96.6% (483/500) if a semi-annual PC is implemented when programmes restart, in moderate endemicity settings. The achievement of the target can be anticipated up to a mean of 4.8 years. The ICL model generates slightly more pessimistic results in relation to the predicted impact of interruptions in PC, on the delay to reach the WHO targets for hookworm, by the two mitigation strategies. This is explained by noting that already in the baseline scenario, the target is reached 0.35 [0; 2] years after the interruption (**Supplementary Table 3**). Both models agree that the two mitigation strategies can accelerate the moment when the 2030 target is reached, by on average more than two years for *A. lumbricoide*s, that is, even a single community-wide round is sufficient to compensate for the year missed in moderate endemicity settings (**Supplementary Table 4**). For all three species and for both models, in high endemicity settings we are unlikely to reach the EPHP target by 2030 even without interruption, according to the current guidelines (less than 5% probability), but a single year of community-wide PC (semi-annual) when programmes restart, will be beneficial for *T. trichiura* in speeding up the progress, according to the EMC model. It shows that in those settings, this mitigation could be essential to increase the probability to reach the target from 7.8% (34/500) to 71.4% (357/500). The ICL model results are in reasonable agreement in showing the beneficial effect of community-wide mitigation.

## Discussion

The objective of this study was to investigate the impact of disruptions of STH control programmes by the COVID-19 pandemic on the progress towards reaching the 2030 morbidity target, and to estimate to what extent that impact can be reduced by mitigation strategies. Two different, and independently developed, stochastic individual-based models of parasite transmission and PC impact were employed to examine these questions. We assumed that the programmes resume after varying periods of interruption up to 18 months in lengths, in order to account for the uncertainty about the duration of the COVID-19 pandemic and related control measures. We found that for all STH species, M&HI prevalence catches up before 2030 in both moderate and high endemicity settings, even without implementing a mitigation strategy: catch-up times are limited to 1 – 3 years (moderate endemicity setting) and 1 – 5 years (high endemicity setting). Mitigation strategies reduce the catch-up time by less than one year on average and at most two years. In some case however, they have the benefit of increasing the probability to reach the target by 2030 or reaching it earlier. Thus, mitigation strategies can be an opportunity to enhance progress towards EPHP, especially when reaching the target is not feasible by 2030.

Our results further show that it will always be advantageous to recover the last round missed as soon as possible, e.g. six months after the initially planned date. Our analyses show that it is feasible to reach the target in moderate endemicity contexts where hookworm or *A. lumbricoides* are the dominant species. Different durations of interruption do not have a strong impact on reaching the target but they can introduce a delay up to 1.5 [0; 3] years (ICL, hookworm) and up to 1.4 [0; 3] years (EMC, *A. lumbricoides*).

Although the mitigation strategies considered here have a limited impact on catch-up times, they will help to accelerate the achievement of EPHP by 2030 in moderately *A. lumbricoides* endemic settings. A single community-wide round would therefore be suggested to compensate for a missed year of PC. Mitigation strategies will not help to increase the low probability of reaching the target in the context of *T. trichiura*, but they will enhance the progress by lowering the prevalence of M&HI infections. Both models agree that in highly endemic settings for all three species, the target will not be reached by 2030, even without interruption of PC. For *T. trichiura*, this is mainly due to the relatively low efficacy (60%) of albendazole treatment; dual treatment with ivermectin would be required to reach the target.^9^ For high endemicity areas, adding a year of semi-annual community-wide PC (two rounds) as mitigation, proves to be crucial to speed up the progress and enhance the chance to reach the target for *A. lumbricoides* (ICL) and *T. trichiura* (EMC).

The discrepancies observed in the results between the two models can be explained by different assumptions, such as regarding age patterns in exposure to eggs/larvae in the environment. For instance, the ICL model assumes for hookworm a flat age profile making the exposure to the infection independent of age. In contrast, in the EMC model it is assumed that exposure increases during the first ten years of life and then stabilises, such that most of the hookworm infection is carried by adults and higher worm burden is observed in adults than in SAC. The two different assumptions about age pattern in exposure, fit previously published age-intensity profiles observed in the field,^10,11^ and thus potentially reflect different transmission settings. This difference in assumptions is directly reflected in our results: the probability of reaching the morbidity target in areas with moderate prevalence of hookworm infections is higher in the ICL model, given the current school-based treatment guidelines.^6^

COVID-19-related interruptions are affecting several countries with different histories of control. In this study, we decided to implement the first year of treatment for all STH to simulate ongoing PC; the second year is then missed due to COVID-19 preventive measures. We do not investigate the impact of skipping later rounds of PC. However, theoretically, missing PC rounds early in the programme would have a more detrimental impact on current progress and goals to be achieved. As such, our predictions provide a conservative (i.e. pessimistic) foundation on which to base policy decisions.

The estimated outcomes of this study can be tested in settings where data of M&HI prevalence was collected before the interruption and where the collection will continue after. The Geshiyaro project^12^ and the DeWorm3^13^ focus on the feasibility of interrupting the transmission of parasitic worms by repeated rounds of PC, included STH. In these projects, PC rounds have been delayed by COVID-19 hence they will be a good test of model predictions. Another test of the outcomes proposed with this paper, would require in settings where mitigation strategies are implemented, the assessment if they reach the 2030 target sooner than settings where mitigation strategies have not been implemented. Testable model outcomes fulfil one of the criteria requested by the Policy-Relevant Items for Reporting Models in Epidemiology of Neglected Tropical Diseases (PRIME-NTD) (**Supplementary Table 2**).

Even though STH control programmes may not take long to catch up after the interruption, it is important to minimise the catch-up time and consider mitigation strategies as soon as possible. STH-related morbidity is indeed associated with M&HI infections,^1^ and prolonged interruptions could cause a period of higher M&HI prevalence, long enough to develop morbidity. Anaemia, for example, is the main hookworm-related morbidity and it is strongly associated with moderate and heavy hookworm infections. Anthelminthic treatment is an effective means of improving haemoglobin levels.^14^ A consistent drop in haemoglobin below the WHO threshold defined for anaemia is estimated to occur starting from 2000 eggs per gram (epg), the value defining moderate infections.^15^ It could be, therefore, crucial especially for children, to prevent prolonged periods with high M&HI.

We show that even a single year of community-wide treatment after the interruption speeds up the progress towards the morbidity target. In some cases, the target may even be reached sooner than without interruption. Even though community-wide treatment will require extending drug donations to adults as well as children, in specific settings which are far from reaching the target, this can be, not only helpful in speeding up the process, but also offer an opportunity to reach the morbidity target by 2030.

## Conclusions

We estimated that the COVID-19-related interruption of STH control programmes will only temporarily impact the progress towards the EPHP target by 2030, since for all STH species M&HI prevalence after interruption catch up with the prevalence in the scenarios without interruption, recovering the ground lost by 2030. However, after a PC interruption programmes require a catch-up time (estimated to be less than three years in moderate endemicity settings and less than five years in high endemicity settings) during which endemic areas could attain higher levels of moderate and high infections. We suggest, therefore, to minimise the time without PC and to restart PC as soon as possible, even if that is before the time when the next round of PC would have been scheduled under normal circumstances. In addition, disruption by COVID-19 could be turned into an opportunity to increase the probability of reaching the target in those settings where it is not feasible with the current guidelines, by implementing suitable mitigation strategies. A one-year period of community-wide treatment after the interruption would speed up the progress towards the morbidity target. In some cases, resuming programmes with a mitigation strategy would be the only possibility to reach the morbidity target by 2030.

## Data Availability

The simulations results of this article are available from the corresponding author on request.

## Authors’ contributions

VM, FG, CV contributed to concept and design. VM, FG, CV, SB, KMM contributed to the analyses. All authors contributed to data interpretation. VM, FG contributed to drafting of manuscript. All authors contributed to critical review and revision of manuscript. All authors read and approved the final manuscript. FG is the guarantor of the paper.

## Acknowledgements

The authors would like to thank all of the NTD Modelling Consortium members.

## Ethical approval

Not required.

## Data Availability Statement

The simulations’ results of this article will be shared on reasonable request to the corresponding author.

