## Supplementary material for "Modelling the impact of COVID-19-related control programme interruptions on progress towards the WHO 2030 target for soil-transmitted helminths": Supplementary Figure 1.docx

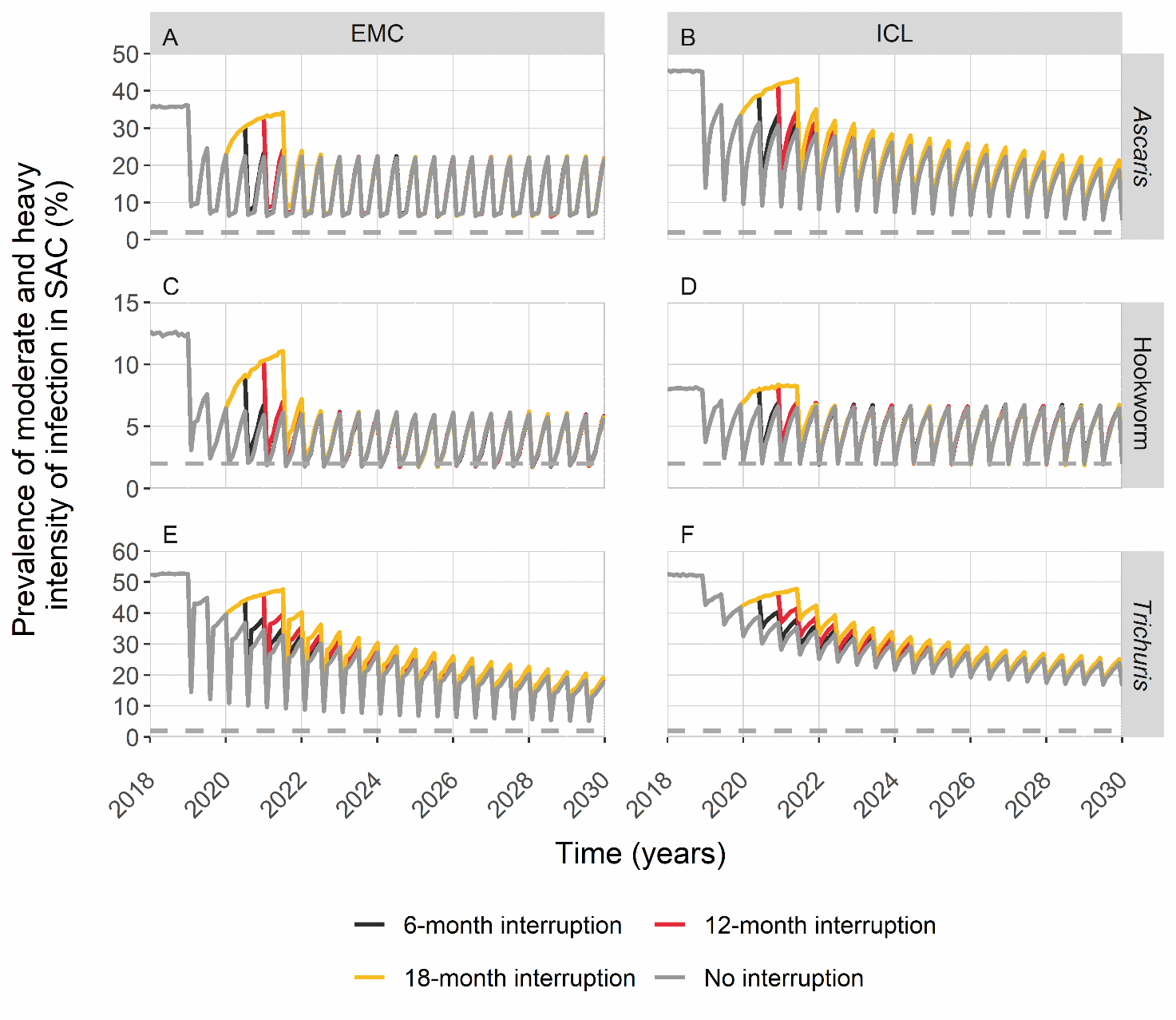

**Supplementary Figure 1.** Timeline of moderate-to-heavy intensity infections prevalence in high endemicity settings, if programmes resume without mitigation strategies. The comparison between “no interruption” and restarting after 6, 12 or 18 months is presented by line colours. The horizontal dashed line represents the 2% threshold set by the WHO to assess the goal. Results from both the EMC model (A, C, E panels) and the ICL model (B, D, F panels) are shown, for all STH species.
