## Supplementary material for "Modelling the impact of COVID-19-related control programme interruptions on progress towards the WHO 2030 target for soil-transmitted helminths": Supplementary Figure 2.docx

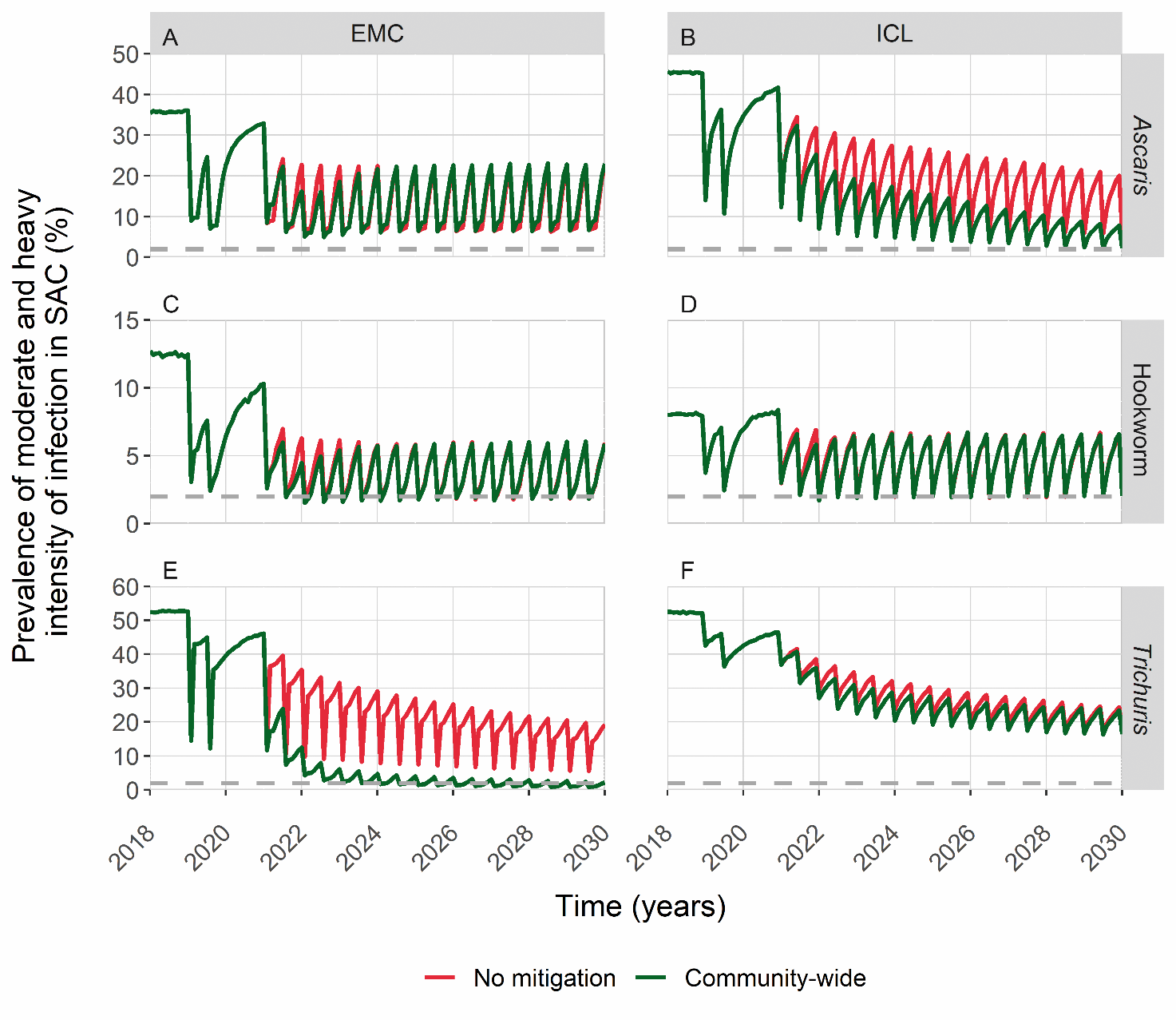

**Supplementary Figure 2.** Timeline of moderate-to-heavy intensity infections prevalence in high endemicity settings, if programmes resume one year after the interruption, i) without mitigation strategies (red line), ii) providing a first year of community-wide MDA before going back to the current WHO treatment guidelines (dark green line). The horizontal dashed line represents the 2% threshold set by the WHO to assess the goal. Results from both the EMC model (A, C, E panels) and the ICL model (B, D, F panels) are shown, for all STH species.
