## Supplementary material for "Modelling the impact of COVID-19-related control programme interruptions on progress towards the WHO 2030 target for soil-transmitted helminths": Supplementary Table 1.docx

Supplementary Table 1. Model parameters used to simulate transmission of *Ascaris lumbricoides*, *Trichuris trichiura* and hookworm infections.

|  | **Value or assumption** | |
| --- | --- | --- |
| **Parameter** | **Erasmus MC** | **Imperial College London** |
| **Human demography** | | |
| *Hookworm* | Demographic data quantified for sub-Saharan Africa 2000 United Nations Population Division.^1^ | Demographic data taken from 2003 Kenya Demographic and Health Surveys.^2^ |
| *Ascariasis* | Indian fertility and mortality rates as reported for 1980-1985 by United Nations Population Division (2015 Revision). | Demographic data taken from 2003 Kenya Demographic and Health Surveys.^2^ |
| *Trichuris* | Indian fertility and mortality rates as reported for 1980-1985 by United Nations Population Division (2015 Revision). | Demographic data taken from 2003 Kenya Demographic and Health Surveys.^2^ |
| **Transmission of infection** | | |
| Seasonal variation in contribution to reservoir | Stable throughout the year (assumption). | Stable throughout the year (assumption). |
| Aggregation of parasites in hosts |  |  |
| *Hookworm* | $k_{w}=0.35$ ^2^ | $k_{w}=0.35$ ^2^. |
| *Ascariasis* | $k_{w}=0.8$ ^3^ | $k_{w}=0.8$ ^3,4^ in high-prevalence settings,  $k_{w}=0.2$ in moderate-prevalence settings. |
| *Trichuris* | $k_{w}=0.38$, in high-prevalence settings fitted to data^5^,  $k_{w}=0.12$ in moderate-prevalence settings. | $k_{w}=0.38$, in high-prevalence settings fitted to data^5^,  $k_{w}=0.12$ in moderate-prevalence settings. |
| Variation in exposure and contribution to the environmental reservoir by age and sex |  |  |
| *Hookworm* | Relative exposure and contribution to the reservoir both increase linearly from 0 to 1 between ages 0–10 and is stable thereafter with no difference between males and females.^6^ | Relative exposure and contribution to the reservoir are assumed to vary be constant across age groups, assuming no difference between males and females. The values were estimated from baseline data of the Tumikia study^7^ and unpublished epidemiological data of the DeWorm3 study. |
| *Ascariasis* | Contribution to the reservoir increases linearly from 0 to 1 between ages 0–10 and is stable thereafter with no difference between males and females (reflecting behaviour related to defaecation and mobility patterns as previously estimated for hookworm^6^). Exposure to the reservoir is defined as a piece-wise linear function of age that increases linearly from a base level $x_{0}$ =0.33 of relative exposure at age zero to a relative exposure of 1.0 at age $a_{\text{peak}}$=3, and then again linearly declines back to the base level $x_{0}$ at age 15 and is stable thereafter. This function aims to reflect behaviour leading to ingestion of contaminated matter, which typically peaks in young children.^3^ | Relative exposure and contribution to the reservoir by age are assumed to be equal and are estimated from the baseline data: 0.22 (0-4 years), 1.88 (5-9), 1.0 (10-19), 0.53 (20+). |
| *Trichuris* | Contribution to the reservoir increases linearly from 0 to 1 between ages 0–10 and is stable thereafter with no difference between males and females (reflecting behaviour related to defaecation and mobility patterns as previously estimated for hookworm^6^). Exposure to the reservoir is defined as a piece-wise linear function of age that increases linearly from a base level $x_{0}$ =0.33 of relative exposure at age zero to a relative exposure of 1.0 at age $a_{\text{peak}}$=3, and then again linearly declines back to the base level $x_{0}$ at age 15 and is stable thereafter. This function aims to reflect behaviour leading to ingestion of contaminated matter, which typically peaks in young children.^3^ | Relative exposure and contribution to the reservoir are assumed to vary piece-wise constant by age group and are estimated at 0.3 (0-4 years), 1.28 (5-14), 1 (15-24) and 0.17 (ages 25+), assuming no difference between males and females. These figures were estimated from epidemiological data.^5^ |
| **Life history and productivity of the parasite in the human host** | | |
| Average worm lifespan |  |  |
| *Hookworm* | 3 years ^8, 9, 10^ | 2 years ^11^ |
| *Ascariasis* | 1 year ^3,8, 9, 10,12^ | 1 year ^8,9,10,12^ |
| *Trichuris* | 1 year ^5,11^ | 1 year ^5,11^ |
| Variation in worm lifespan | Weibull distribution with shape 2; i.e. the mortality rate is zero at age zero and then increases linearly with worm age (assumption as previously used for hookworm ^6^). | Exponential distribution; i.e. the mortality rate is constant and independent of worm age. |
| Pre-patent period |  |  |
| *Hookworm* | 7 weeks ^8,9,13,14^ | No pre-patent period used. |
| *Ascariasis* | 10 weeks ^8^ | No pre-patent period used. |
| *Trichuris* | 10 weeks ^8^ | No pre-patent period used. |
| Age-dependent reproductive capacity | Constant over age (assumption). | Constant over age (assumption). |
| Female worm fecundity | Density-dependent on total number of female worms in host, assuming hyperbolic saturation.^6^ | Density-dependent on total number of female worms in host, assuming exponential saturation. Exponential model of saturation with parameter γ = 0.02 ^15^ for hookworm, γ = 0.07 for ascaris ^4^ and γ = 0.0035.^5,16^ |
| *Hookworm* | On average 8.3 eggs per female worm per 41.7 mg sample of faeces (200 epg per female worm, as previously reported based on association between number of expulsed adult female worms and egg counts based on Kato-Katz^17^). The average maximum total host output is assumed to be 62.5 eggs per 41.7 mg faeces (1500 epg, as previously assumed^6^). | On average 3 eggs per female worm per 41.7 mg sample of faeces (72 epg per female worm, as previously reported based on association between number of expulsed adult female worms and egg counts based on Kato-Katz^17^). |
| *Ascariasis* | On average 406 eggs per female worm per 41.7 mg sample of faeces (9750 epg per female worm), and maximum total host output of 777 eggs per 41.7 mg faeces on average (18,650 epg). These figures were estimated from pre-control data on number of expulsed adult female worms and egg counts based on a concentration and sedimentation technique using homogenised stools.^3^ | On average 320 eggs per female worm per 41.7 mg sample of faeces (7674 epg per female worm). |
| *Trichuris* | On average 15.4 eggs per female worm per 41.7 mg sample of faeces (370 epg per female worm), and maximum total host output of 3333.33 eggs per 41.7 mg faeces on average (80,000 epg). These figures were estimated from pre-control data on number of expulsed adult female worms and egg counts based on a concentration and sedimentation technique using homogenised stools.^3^ | On average 5.875 eggs per female worm per 41.7 mg sample of faeces (141 epg per female worm).^5^ |
| Host immunity to incoming infections | None (assumption). | None (assumption). |
| **Infection dynamics in environmental reservoir** | | |
| Survival of infective material in the central reservoir | Exponential survival (assumption). | Exponential survival (assumption). |
| *Hookworm* | Average lifespan of two weeks, implemented as a monthly survival probability of $\exp\left( -26/12 \right)=11.5\%$ (95%-CI: 0.05–7.38 weeks under assumption of exponential survival), based on the notion that average survival time is in the order of weeks.^13,14,18^ | Average lifespan of 30 days.^11^ |
| *Ascariasis* | Average lifespan of 1.5 month, implemented as a monthly survival probability of $\exp\left( -1/1.5 \right)=51.3\%$ (95%-CI: 0.04–5.53 months under assumption of exponential survival).^9,10^ | Lifespan of approximately 2 months.^19^ |
| *Trichuris* | Average lifespan of 20 days implemented as a monthly survival probability of $\exp\left( -1/(2/ 3 \right))=22.3\%$ (95%-CI: 0.02–2.46 months under assumption of exponential survival. | Lifespan of approximately 20 days.^5^ |
| **Drug treatment** | | |
| Proportion of adult worms killed by single dose of albendazole (400 mg), or pyrantel pamoate (10 mg/kg, ascariasis only) | Assumption: proportion killed is equal to the faecal egg reduction rate. | Assumption: proportion killed is equal to the faecal egg reduction rate. |
| *Hookworm* | 0.95 for albendazole.^20^ | 0.95 for albendazole.^20^ |
| *Ascariasis* | 0.99 for albendazole.^20^ | 0.99 for albendazole.^20^ |
| *Trichuris* | 0.60 for albendazole.^20^ | 0.60 for albendazole.^20^ |
| **Diagnostic test outcomes** |  |  |
| Variability in measured host load of infective material (eggs per examined sample of faeces) |  |  |
| *Hookworm* | Kato-Katz: negative binomial distribution with aggregation parameter $k=0.35$, estimated separately from repeated individual-level egg count data from Uganda.^21^ | Kato-Katz: negative binomial distribution with aggregation parameter $k=0.35$, estimated from unpublished triple egg count data from Tamil Nadu, India |
| *Ascariasis* | Kato-Katz: negative binomial distribution with aggregation parameter $k=0.25$. | Kato-Katz: negative binomial distribution with aggregation parameter $k=0.3$^22^ |
| *Trichuris* | Kato-Katz: negative binomial distribution with aggregation parameter $k=0.82$. | Kato-Katz: negative binomial distribution with aggregation parameter $k=0.82$^16^ |
| Cut-offs for no, light, moderate, and heavy infection |  |  |
| *Hookworm* | 1, 2000, and 4000 epg | 1, 2000, and 4000 epg |
| *Ascariasis* | 1, 5000, and 50,000 epg | 1, 5000, and 50,000 epg |
| *Trichuris* | 1, 1000, and 10,000 epg | 1, 1000, and 10,000 epg |
