## Supplementary material for "Modelling the impact of COVID-19-related control programme interruptions on progress towards the WHO 2030 target for soil-transmitted helminths": Supplementary Table 2.docx

**Supplementary Table 2:** The Policy-Relevant Items for Reporting Models in Epidemiology of Neglected Tropical Diseases (PRIME-NTD).

| **Principle** | **What has been done to satisfy the principle?** | **Where in the manuscript is this described?** |
| --- | --- | --- |
| **Stakeholder engagement** | Work has been presented at the following WHO webinars: (i) Neglected Tropical Diseases and COVID-19: Impact on Programme Implementation; and (ii) A Research Agenda for NTD Programmes Affected by the COVID-19 Pandemic. | - |
| **Complete model documentation** | The scenarios modelled in this study are described in the manuscript. Transmission models used are described in the manuscript and fully documented in previous publications.^1,2,3^ | Methods section and References. |
| **Complete description of data used** | Data and parameters used are described in the manuscript and in the supplementary information. | Methods section and Supplementary Table 1. |
| **Communicating uncertainty** | We consider stochastic uncertainty in communicated results and we account for quantitative differences between the two models used. | Methods, Results and Discussion sections. |
| **Testable model outcomes** | The model outcomes can be tested by collecting data once programmes resume. | Discussion section. |
