## Supplementary material for "Modelling the impact of COVID-19-related control programme interruptions on progress towards the WHO 2030 target for soil-transmitted helminths": Supplementary Table 3.docx

**Supplementary Table 3.** Time (years since the interruption of the control programme) and probability to reach the 2030 target in the baseline scenario (no interruption); introduced delays (years) in reaching the target for each interruption scenario, compared to the baseline scenario. Hookworm, moderate endemicity setting.

| **Model** | **Time (mean [95%CI]) and probability to reach the target in the baseline scenario** | **Delay from the baseline scenario**  **(mean [95% CI]) (ys)** | | | | |
| --- | --- | --- | --- | --- | --- | --- |
|  |  | **6-month interruption** | **12-month interruption** | **18-month interruption** | **Semi-annual mitigation** | **Community-wide mitigation** |
| EMC | 8.14 [1; 10]  66.6% | 0.34 [-2; 2] | 0.17 [-2; 2] | 0.49 [-2; 2] | -4.76 [-8; -2] | -1.32 [-3; 0] |
| ICL | 0.35 [0; 2]  100% | 0.39 [-1; 1] | 1.27 [0; 3] | 1.49 [0; 3] | 1.17 [0; 2] | 1.19 [0; 2] |
