## Supplementary material for "Modelling the impact of COVID-19-related control programme interruptions on progress towards the WHO 2030 target for soil-transmitted helminths": Supplementary Table 4.docx

**Supplementary Table 4.** Time (years since the interruption of the control programme) and probability to reach the 2030 target in the baseline scenario (no interruption); delays (years) in reaching the target for each interruption scenario, compared to the baseline scenario. *A. lumbricoides*, moderate endemicity setting.

| **Model** | **Time (mean [95%CI]) and probability to reach the target in the baseline scenario** | **Delay from the baseline scenario**  **(mean [95% CI]) (ys)** | | | | |
| --- | --- | --- | --- | --- | --- | --- |
|  |  | **6-month interruption** | **12-month interruption** | **18-month interruption** | **Semi-annual mitigation** | **Community-wide mitigation** |
| EMC | 6.37 [2; 10]  83.2% | 0.52 [-1; 2] | 1.35 [0; 2] | 1.40 [0; 3] | -2.30 [-4; -1] | -2.45 [-4; -1] |
| ICL | 7.96 [5, 10]  26.2% | 0.85 [-3; 7] | 1.90 [-2; 9] | 2.44 [-2; 9] | -2.42 [-5; 0] | -1.67 [-5; 2] |
